# Cytomegalovirus latency exacerbates cardiac inflammation and tissue remodeling after myocardial infarction

**DOI:** 10.1101/2025.10.27.25338932

**Authors:** Eleni Dapergola, Marko Šustić, DiyaaElDin Ashour, Maja Cokarić Brdovčak, Jelena Materljan Franki, Mijo Golemac, Berislav Lisnić, Valèrie Boivin-Jahns, Tobias Krammer, Kenz Le Gouge, Lavinia Rech, Verena Stangl, Karl Kashofer, Peter P. Rainer, Roland Jahns, Encarnita Mariotti-Ferrandiz, Maxim Terekhov, Hamid Marwan, Mohammadreza Keshtkar, Antoine-Emmanuel Saliba, Clement Cochain, Stipan Jonjić, Ulrich Hofmann, Stefan Frantz, Ilija Brizić, Georg Gasteiger, Gustavo Campos Ramos

**Affiliations:** Würzburg Institute of Systems Immunology, Max Planck Research Group at the Julius-Maximilians-University Würzburg, Würzburg, Germany; Department of Internal Medicine, Faculty of Medicine, University of Rijeka, Rijeka, Croatia; Department of Internal Medicine I, University Hospital Würzburg, Würzburg, Germany; Comprehensive Heart Failure Center, University Hospital Würzburg, Würzburg, Germany; Center for Proteomics, Faculty of Medicine, University of Rijeka, Rijeka, Croatia; Department of Histology, Faculty of Medicine, University of Rijeka, Rijeka, Croatia; Institute of Pharmacology and Toxicology, University of Würzburg, Würzburg, Germany; Helmholtz Institute for RNA-based Infection Research, Helmholtz Centre for Infection Research, Würzburg, Germany; Division of Cardiology, Department of Internal Medicine, University Heart Center, Medical University of Graz, Graz, Austria BioTechMed Graz, Graz, Austria; Department of Internal Medicine, Landeskrankenhaus Südweststeiermark, Wagna, Austria (V.S), Diagnostic and Research Institute of Pathology, Medical University of Graz, Austria (V.S); Diagnostic and Research Institute of Pathology, Diagnostic and Research Center of Molecular BioMedicine, Medical University of Graz; Immunoregulation-Immunopathology-Immunotherapy, UMRS959, Sorbonne Université, INSERM, Paris, France; Comprehensive Heart Failure Center (CHFC), Department of Molecular and Cellular Imaging, University Hospital Würzburg, Würzburg, Germany; Department of Medicine, St. Johann in Tirol General Hospital, St. Johann in Tirol, Austria; Interdisciplinary Bank of Biomaterials and Data Würzburg (ibdw), University Hospital Würzburg, Würzburg, Germany; Université Paris Cité, Inserm, PARCC, 56 rue Leblanc, 75015 Paris, France; Institute of Experimental Biomedicine, University Hospital Würzburg, Würzburg, Germany

**Keywords:** Myocardial infarction, T cells, CMV, cardiac remodeling Running title: latent CMV infection impacts post-MI repair

## Abstract

**Background:** Epidemiological studies have consistently associated cytomegalovirus (CMV) seropositivity with adverse cardiovascular outcomes. However, the mechanisms by which CMV infection impacts pathophysiological mechanisms in the heart remain poorly understood. In this study, we sought to dissect how latent murine CMV infection impacts cardiac immune cell dynamics at steady-state and during post-myocardial infarction (MI) repair.

**Methods:** Experimental MI studies were conducted in C57BL/6J mice previously infected with murine CMV (MCMV). In situ inflammatory responses were characterized by spectral flow cytometry, bulk and single-cell RNA / T cell receptor sequencing, whereas cardiac function was monitored by echocardiography and magnetic resonance imaging (cMRI). Moreover, we retrospectively assessed the CMV serostatus in a well-characterized patient cohort with longitudinal cMRI data available and performed bulk T cell receptor sequencing on peripheral blood and myocardial samples to identify CMV-specific TCRs.

**Results:** Our findings show that exposure to MCMV induces long-term changes in the cardiac transcriptional profile and alterations in cardiac-resident immune cell populations, including the establishment of virus-specific memory CD8^+^ T cell residency. Compared to infarcted controls, mice previously exposed to MCMV exhibited stronger inflammatory responses, marked by increased CD8^+^ T cell infiltration, and worsened cardiac function following MI. These observations in mice were supported by data from CMV-seropositive MI patients, who harbored CMV-responsive T cells in the heart.

**Conclusions:** Our findings demonstrate that latent CMV infection leads to long-term changes in the cardiac microenvironment, which ultimately impair post-MI healing outcomes.

## Introduction

It is now widely recognized that immunological processes play key roles in a broad range of myocardial diseases, including post-myocardial infarction (MI) repair and heart failure (HF)^1^. Tissue immunology is shaped by individual histories of antigenic encounters, and common viral infections may therefore influence pathophysiological mechanisms in the heart. However, the possible links between antiviral immunity and myocardial diseases remain poorly understood. Among the most common viral infections in humans, cytomegalovirus (CMV) shows particularly high seroprevalence, exceeding 60% in elderly populations ^2,3^. These observations are significant because latent CMV infection is considered a major driver of variation in individual immune profiles^4^. Moreover, the age-related rise in CMV seropositivity parallels the increased incidence of MI and HF in elderly^5^. In Germany, for instance, the average age of first MI occurrence is 66.6 years for men and 75.3 years for women, whereas the prevalence of HF increases from less than 1% in individuals aged 45– 55 to over 10% among those in their 80s^6,7^.

CMV is one of the most ubiquitous human herpesviruses and can establish persistent, or even completely latent infection in its host with occasional virus reactivation^8,9^. CMV infection elicits a characteristic CD8^+^ T cell response distinguished by a phenomenon known as memory inflation. In this process, T cells targeting specific viral epitopes gradually expand over time, and in older individuals, can comprise up to 50% of the entire circulating CD8^+^ T cell pool^10^. This leads to a markedly skewed T cell receptor (TCR) repertoire, with the affected cells predominantly exhibiting both terminal differentiation phenotype and cellular senescence features^11^. Moreover, latent CMV infection generates chronic low-level inflammation characterized by increased levels of circulating cytokines such as IFN-γ, TNF, and IL-6^5,12^. Not only are these cytokines elevated in patients with chronic heart failure, but IFN-γ also induces detrimental derangements in cardiomyocyte metabolism as characterized by a decreased oxidative phosphorylation rate^13,14^, a phenotype associated with a failing heart^15^. Four decades ago, Adam et al. reported an association between CMV infection and atherosclerotic disease, as they observed higher CMV IgG antibody levels in patients undergoing vascular surgery compared to matched controls^16^. Subsequently, several large epidemiological studies have explored the connections between CMV serostatus and the incidence and prognosis of cardiovascular disease (CVD)^17–20^, and a recent meta-analysis concluded that CMV-positive individuals have a higher risk for cardiovascular mortality^21^. Furthermore, CMV was the single viral agent found most often in the cardiac tissue of patients with fulminant myocarditis^22^, and although it seldom causes the disease itself^23^, the presence of CMV in the cardiac tissue of patients who died from myocarditis indicates its potential contribution to pathophysiological mechanisms in the heart. Hence, multiple lines of evidence suggest that latent CMV infection and the chronic inflammatory environment that it generates, modulate cardiac outcomes in different pathological states.

Despite overwhelming clinical evidence for the association of CMV infections with myocardial diseases, the putative mechanisms underlying this link remain elusive. In this study, we investigated immunological processes during MI superimposed with experimental murine models of viral infections that are common in humans, including murine cytomegalovirus (MCMV), to recapitulate real-life clinical scenarios and to mechanistically dissect how viral infections impact myocardial homeostasis and post-MI inflammation. Our findings reveal that exposure to MCMV led to the establishment of virus-specific T cells in cardiac tissue, which persisted long after the acute infection resolved and transitioned into latency. Through a combination of cellular and functional techniques, including spectral flow cytometry, single-cell RNA / TCR sequencing, echocardiography, and cardiac magnetic resonance imaging (cMRI), we found that latent MCMV infection is associated with profound changes in the cardiac tissue-resident immune cell compartment, increased post-MI inflammatory responses, and pronounced adverse cardiac remodeling. Finally, analysis of an MI patient cohort stratified by CMV serostatus and longitudinal cMRI demonstrated that adaptive immune responses to CMV correlate with worsened cardiac function. Collectively, these findings provide initial evidence that CMV seropositivity may serve as a clinical marker to predict post-MI healing outcomes and cardiac remodeling in MI patients, thus highlighting novel avenues for stratifying patients according to their expected inflammatory burden.

## Methods

### Data availability

The detailed methods and additional figures are available in the Supplemental Material. Transcriptomic datasets from this study have been archived in NCBI’s Gene Expression Omnibus (GEO) database (GSE309570 and GSE309706). Animal procedures were approved by the local authorities (Regierung von Unterfranken and Animal Welfare Committee at the University of Rijeka) and conformed to the guidelines from Directive 2010/63/EU of the European Parliament on the protection of animals used for scientific purposes. For experiments conducted in Würzburg, male and female C57BL/6J mice were purchased from Charles River Laboratories and were kept in individually ventilated cages under pathogen-free conditions in animal facilities. For the sequential infections mice were used at the age of four weeks, while for the procedures with MCMV and MI, mice were used after the age of eight weeks. All animals were kept under a 12h-12h light-dark cycle at 22°C and relative humidity (40–50%) with ad libitum access to normal food pellets and sterile drinking water. For experiments conducted in Rijeka, adult (6-12 weeks old) female wild-type C57BL/6J mice (strain #000664) were used in experiments. All mice were housed and bred under the conditions mentioned above at the animal facility of the Faculty of Medicine, University of Rijeka, The Animal Welfare Committee at the University of Rijeka, Faculty of Medicine, and The National Ethics Committee for the Protection of Animals Used for Scientific Purposes (Ministry of Agriculture) approved all animal experiments. The use of human tissue samples from Graz is conformed with legal and institutional requirements and was approved by the ethics committee of the Medical University of Graz (31-288 ex 18/19). The ETiCS study from Würzburg was approved by the ethics commission of the University of Würzburg (186/09) and met the criteria of the Declaration of Helsinki. All patients provided written informed consent.

### Statistical analysis

All data are depicted as the mean ±SEM together with the distribution of individual values for each group. Sample sizes as well as tests used for each statistical analysis are mentioned in the figure legends. Both graphs and statistical analyses were performed using GraphPad Prism version 10.6.1 for Windows (GraphPad Software, Boston, Massachusetts USA, www.graphpad.com) and R studio. To compare 2 groups with data following normal distribution a 2-tailed *t*-test was performed. For multiple comparisons including more than 2 groups an ordinary one-way ANOVA was performed. For non-parametric or mixed data a Tukey’s multiple comparisons test was conducted. A *P* value less than 0,05 was considered statistically significant.

## Results

### Cytomegalovirus infection persistently alters the cardiac immunological landscape

Previous studies conducted both on post-mortem human tissue^22^ and experimental mouse models^24^ have detected CMV in the hearts of infected mammals. To determine the kinetics of viral replication in the heart, we infected C57BL/6J mice with murine CMV (MCMV) and performed a plaque-forming assay on cardiac tissue at different time points (Fig. 1a). As shown in Fig. 1b, viral titers peaked at day 5 post infection (p.i.) and subsequently declined, with no in situ replication detected on day 12 p.i. (Fig. 1b). Additionally, using immunohistochemistry, we identified clusters of cigar-shaped infected nuclei at day 5 p.i. (Fig. 1c). These findings confirm that MCMV can infect murine hearts, yet potential long-term consequences of this infection have not been dissected in detail. CMV usually causes a brief, mild acute infection but maintains lifelong latency. To this end, we investigated the extent of transcriptional changes in the hearts of infected animals during the latent phase of infection. We performed bulk RNA sequencing (RNA-seq) of cardiac tissue from mock, acutely, and latently infected mice. Principal component analysis (PCA) plots demonstrated clear separation of the experimental conditions, suggesting distinct transcriptional states of the cardiac tissue in uninfected, as well as in acutely and latently infected animals (Fig. 1d). Indeed, when comparing gene expression patterns in hearts from latently infected versus non-infected animals, using a stringent cut-off (p_adj_ < 0.01), we detected more than 500 differentially expressed genes (Fig. 1e). Subsequent gene ontology over-representation analysis revealed that the latent MCMV infection caused significant changes in the immune landscape within cardiac tissue. A significant proportion of the overrepresented terms with predominantly upregulated genes was related to inflammatory response, and included categories such as “cell killing”, “positive regulation of T cell activation” and “cellular response to interferon beta” (Fig. 1f). Furthermore, closer inspection of deregulated genes revealed that, compared to uninfected animals, latent CMV infection caused increased expression of not only numerous interferon-related genes, chemokines, and the components of antigen-presenting machinery but also genes associated with cytotoxic T cell response and CCR2^+^ macrophages, such as *Gzma, Gzmb, Cd8, Cd3* and *Ccr2* (Fig. 1g). Interestingly, over-represented GO categories with predominantly down-regulated genes were related to ATP synthesis and oxidative phosphorylation (Fig. 1f and 1g).

**Fig. 1.**
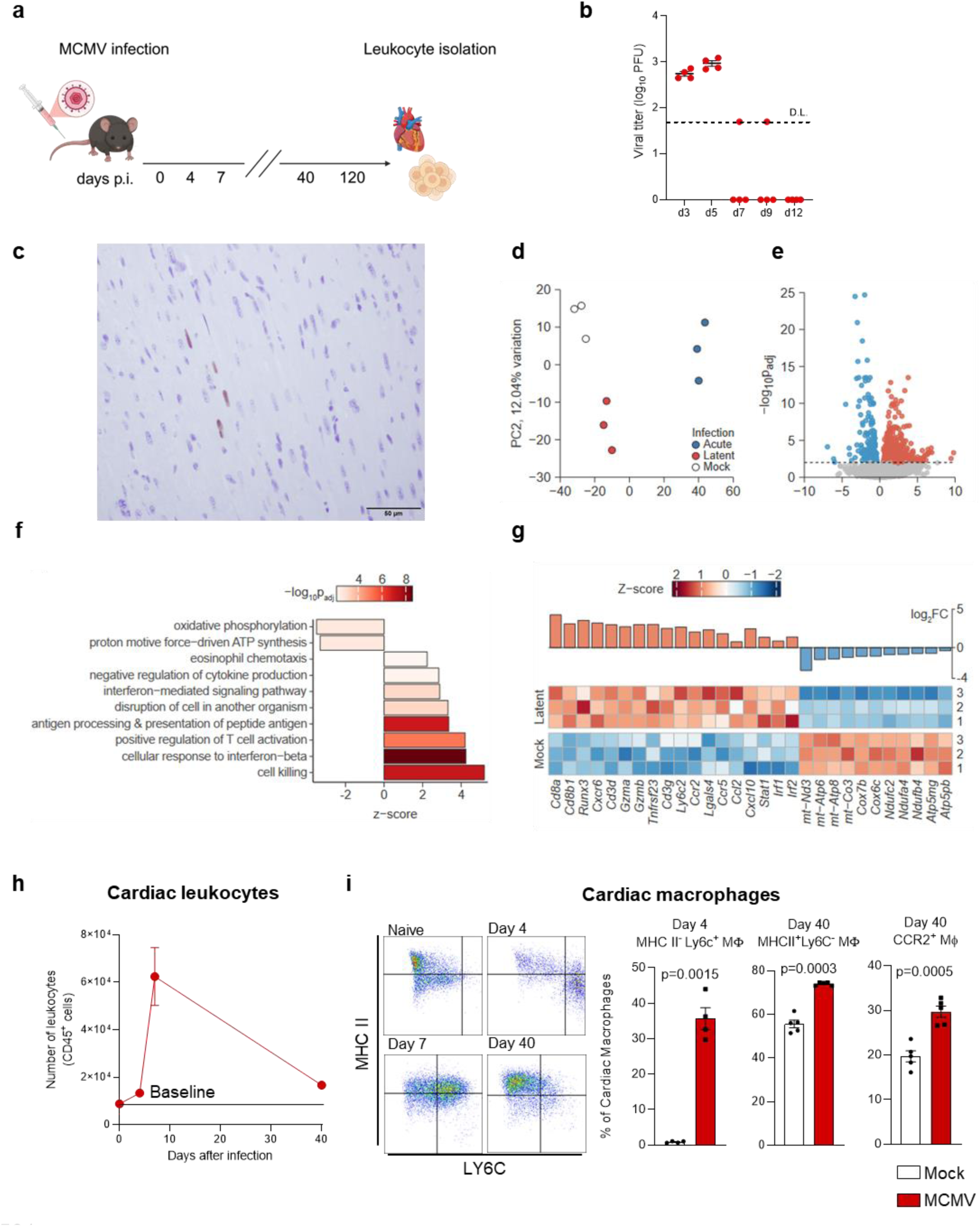
Cytomegalovirus infection persistently alters the cardiac immunological landscape. **a.** Scheme. Mice were infected i.v. with 2×10^5^ PFU of Δm157 MCMV. Non-infected animals were mock treated with the same volume of sterile PBS. **b.** Viral titers, as determined by viral plaque assay, in the hearts of infected mice, at indicated time points (n=4/time point). **c.** Immunohistochemistry staining with IE-1 antibody (magnification: left 10x, right 40x). Mice were sacrificed at day 5 post infection. Brown nuclei represent MCMV-infected cells. **d.** PCA plot showing the clustering of RNA transcriptomic data from non-infected, acutely infected (day 5), and latently infected (day 56) mice. **e, f, g.** Differentially expressed genes and over-represented GO terms in latently infected vs. non-infected animals. **h.** Leukocyte numbers as determined by flow cytometry in the hearts of MCMV-infected mice at indicated time points after infection. The baseline level is set at the number of cardiac leukocytes in non-infected animals. **i.** Representative plots and graphs indicating the phenotypic changes in cardiac monocyte/macrophage populations at indicated time points after infection. Genes with padj<0.01 were considered statistically significantly differentially expressed (**e, f, g**). Statistical analysis (**i**) was performed using an unpaired Student t-test, and exact p values are indicated. Experiments in **h-i** were repeated at least two times. Viral titration (**b**) and RNA sequencing ^50^ were each performed once.

Because the data revealed major changes in cardiac leukocyte composition, we next performed flow cytometry-based phenotyping of cardiac leukocytes (CD45^+^ cells). Leukocyte numbers rose sharply during acute infection, subsequently diminished as the acute phase of the infection ended, but remained approximately twofold greater during latent infection (day 40) compared to levels in non-infected animals (Fig. 1h). Further, cardiac monocyte/macrophage populations underwent phenotypic changes during different phases of MCMV infection. There was an initial influx of Ly6C^+^ MHCII^-^ *bona fide* monocytes during the first several days of infection. At later timepoints, the majority of cardiac monocytic cells expressed MHCII molecules and, and progressively lost Ly6C expression (Fig. 1i). These findings demonstrate that CMV triggers important shifts in the myocardial microenvironment, marked by elevated expression of inflammation-associated genes and persistent shifts in the cardiac-resident immune cell composition.

### Cytomegalovirus infection promotes seeding of tissue-resident CD8^+^ T cells in the myocardium

Next, we characterized the cardiac CD8^+^ T cells enriched upon MCMV infection to determine their antigen specificity and phenotype. The number of cardiac CD8^+^ T cells peaked at day 7 (p.i.) and remained increased even on day 120 p.i. (Fig. 2a). Importantly, the majority of cardiac CD8^+^ T cells at day 120 exhibited an antigen-experienced phenotype, as indicated by CD44 expression (Fig 2b). To determine whether these effects were related to CMV infection or reflected immune alterations induced in response to other viral infections as well, we sequentially infected mice with MHV-68, a murine model of the human pathogen EBV, followed by MCMV and the WSN strain of Influenza A virus, thereby modeling a series of mild infections that resemble the diverse microbial exposures encountered by humans^25^. Seven months after the last infection, we evaluated the presence of virus-specific CD8^+^ T cells in the hearts and lungs by staining with tetramers loaded with peptide antigens of the indicated viruses (Fig. S1A-C). While Influenza and MHV-68-specific T cells were relatively rare in the heart, which could be related to this sequential infection model, we observed again a clear enrichment of MCMV-specific CD8^+^ T cells, thereby confirming that CMV infection causes the long-term persistence of virus-specific cells in the heart also in hosts with multiple exposures to infection. To further characterize these cells, we utilized intravascular CD45 staining as well as the expression of CD69, a surrogate marker of tissue residency, and compared the proportion of CD8^+^ T cells in the heart to that in the lung^26^ (Fig. 2c). FACS analysis showed that a relatively high fraction of CD8^+^ T cells was located extravascularly in the tissue (ivCD45-) and expressed CD69 in the heart, as compared to the lung. We then employed a major histocompatibility complex-I (MHC-I) tetramer specific for the MCMV-derived m139 epitope to identify inflationary antiviral memory CD8^+^ T cells, which expand and persist during MCMV latency^27^. Compared to lungs, a higher proportion of m139⁺ cells in the heart were extravascular, thus indicating the long-term presence of inflationary memory CD8⁺ T cells within cardiac tissue up to 7 months p.i. regardless of any additional exposure to pathogens (Fig. 2d). Furthermore, the majority of these cardiac CD8⁺ T cells co-expressed markers indicative of tissue residency, including CXCR6, CD69, CD103, and CD38, and lacked markers of the circulating Teff pool, such as CX3CR1 and KLRG1 (Fig. 2e). Taken together, these findings reveal that MCMV infection instigates the enrichment and long-term maintenance of tissue-resident memory CD8⁺ T cells within the heart.

**Fig. 2.**
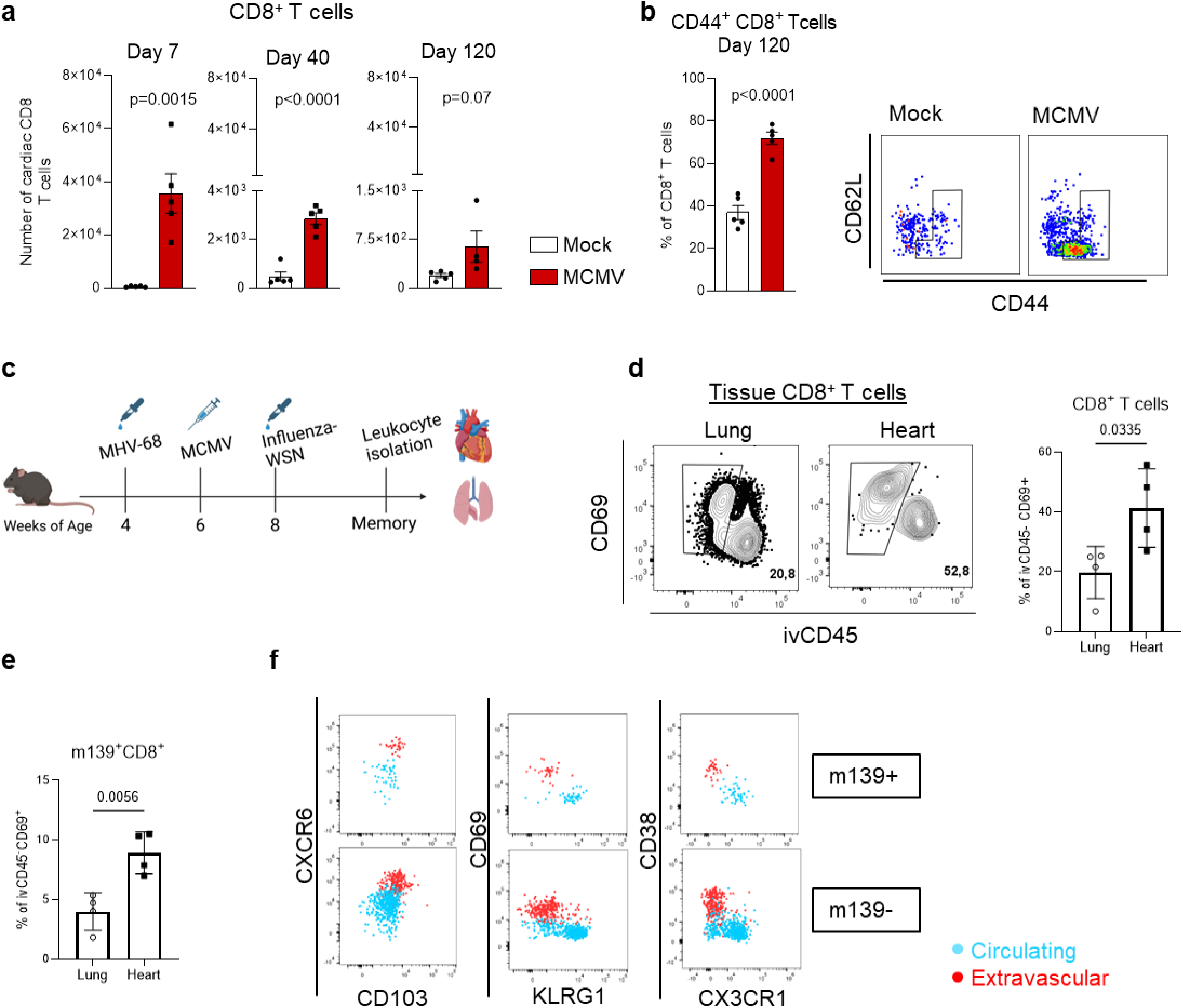
Cytomegalovirus infection promotes seeding of tissue-resident CD8+ T cells into the myocardium. **a.** Cardiac CD8^+^ T cell numbers at indicated time points. **b.** Percentage of antigen-experienced CD8^+^ T cells in the cardiac tissue at day 120 after MCMV infection. **c.** Scheme. Four week old C57BL/J6 mice were sequentially infected intranasally with 10^5^ PFU murine gammaherpesvirus 68 (MHV-68), intraperitoneally with 10^5^ PFU murine cytomegalovirus (MCMV) and intranasally with 10^3^ PFU influenza A/WSN. Hearts and lungs were isolated and analysed 7 months after last infection **d.** Representative FACS plots displaying the gating of extravascular CD8^+^ T cells in the lung and heart 7 months after sequential infections. **e.** Proportion of extravascular M45- and m139-specific cells in the heart (n=4). **f.** Merged representative FACS plots illustrating the tissue-resident (shown in red) or circulating (shown in blue) phenotype of m139-specific and m139-polyclonal CD8^+^ T cells. (**a, b, d, e**). 4-week old female C57BL/6J mice were used in **a** and **b**, while 6–12-week-old females were used in panels **c**–**f**. Statistical analysis was performed using unpaired Student t-test and exact p values are indicated.

### Latent cytomegalovirus drives adverse ventricular remodeling after myocardial infarction in mice

Epidemiological data associate CMV infection with a higher risk of cardiovascular death^21^. Our preliminary data demonstrated that MCMV-infected animals have an enhanced pro-inflammatory transcriptional profile correlating with increased leukocyte infiltration. Therefore, to assess the impact of MCMV infection on cardiac remodeling and function in the context of a noxious event such as myocardial infarction (MI), we subjected latently infected animals and non-infected controls to left anterior descending (LAD) coronary artery ligation (28 days post MCMV infection). Cardiac function was assessed by echocardiography on day 5 post-MI, and hearts were isolated for immunophenotyping (Fig. 3a). While no differences on overall survival were observed (Fig. 3b), infarcted mice previously infected with MCMV showed worsened cardiac function, marked by increased left ventricle end-systolic and - diastolic diameters (Fig. 3c, S2). Moreover, MI superimposed to MCMV infection was associated with decreased fractional shortening at the mid ventricle level (i.e., infarct border zone), compared to MI controls.

**Fig. 3.**
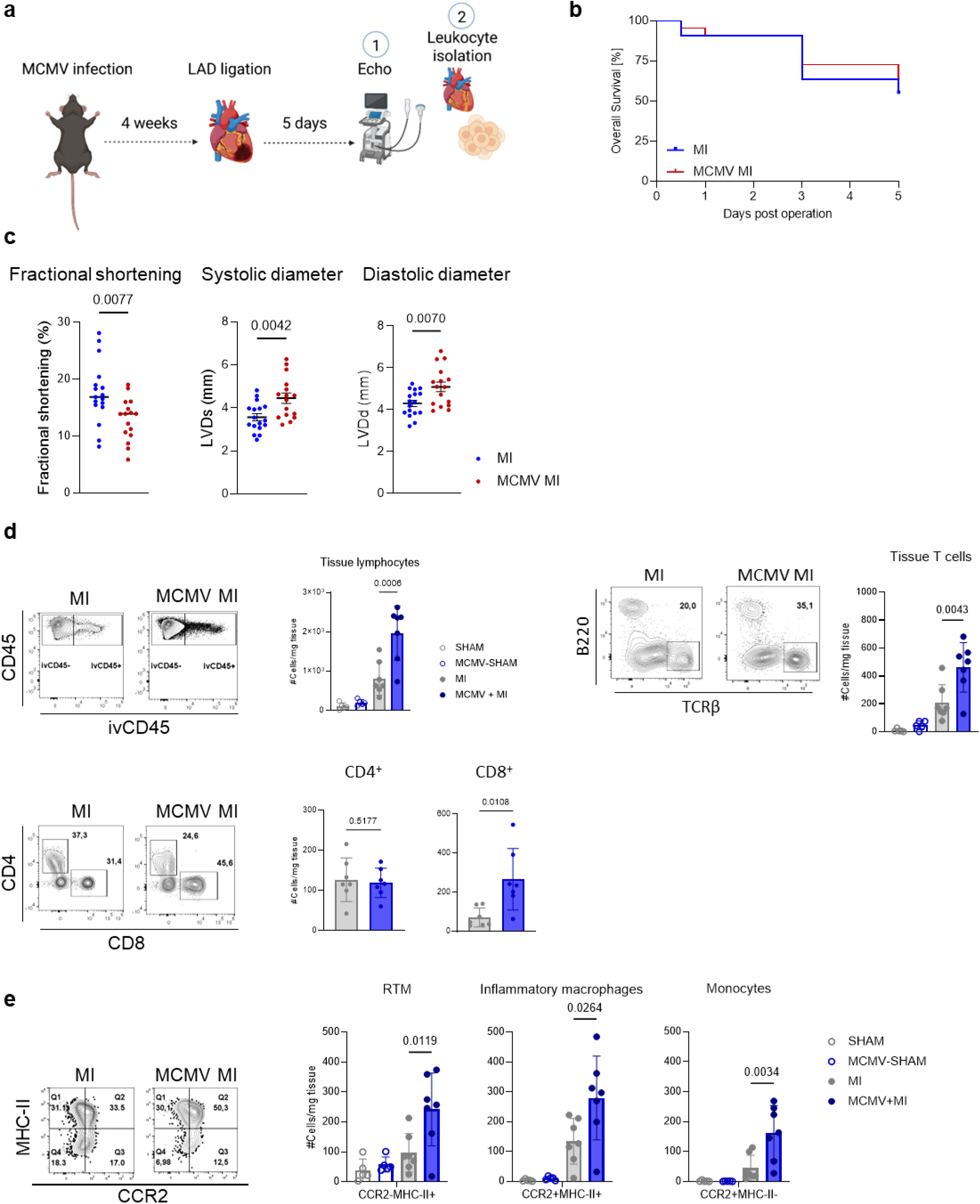
Leukocyte infiltration increases during the acute phase of myocardial infarction in mice with latent cytomegalovirus infection. **a.** Eight-week-old mice were infected with MCMV. At 12 weeks of age, mice underwent LAD ligation. Mice were sacrificed 5 days later and cardiac leukocyte isolation and phenotyping were performed. **b.** Survival after MI. **c.** Fractional shortening of the left ventricle at the mid-papillary level, just above the akinetic area induced by MI. Diastolic and systolic inner diameters of left ventricle as measured by echocardiography. Statistics were calculated using Student t-test; p value is numerically indicated. In MI n=17 and in MCMV MI n=16 **d.** Total cell numbers/mg in heart tissue and representative FACS plots of total tissue lymphocytes and T cells, in SHAM (blank grey), MCMV+SHAM (blank blue), MI (full grey), and MI+ MCMV (full blue) mice cohorts 5 days post MI. **e.** Total cell numbers/mg of RTMs, inflammatory macrophages, and monocytes in heart tissue 5 days post MI. Statistics were calculated using 2-way ANOVA (**d, e**) and Student t-test; for SHAM n=5, for both MI groups n=7. (**c**) p value is numerically indicated.

MCMV infection generates a new steady-state condition in latently infected animals, characterized by chronic inflammatory changes in the pool of heart-infiltrating immune cells, of both lymphoid and myeloid lineages. To further decipher how these immunological shifts impact the response to MI, we conducted a detailed characterization of leukocyte populations infiltrating the infarcted heart. We employed intravascular labeling to distinguish between circulating and tissue-resident immune cells. Cardiac tissue-resident lymphocyte populations were elevated in latently infected and infarcted mice (MCMV+MI) compared to infarcted naive controls. Notably, this increase was primarily driven by an expansion of CD8⁺ T cells in the MCMV+MI group (Fig. 3d). Analysis of infiltrating myeloid populations revealed more abundant tissue resident macrophages (RTMs), inflammatory macrophages, and monocytes in the latently infected infarcted group, as defined by differential expression of CCR2 and MHC-II markers (Fig. 3e). Collectively, these results suggest that a previous/latent MCMV infection exacerbates leukocyte infiltration, predominantly involving CD8⁺ T cells, monocytes and macrophages, into the cardiac tissue during the acute phase post MI.

### Prior MCMV exposure induces an elevated pro-inflammatory transcriptional profile in the cardiac tissue of infarcted mice

To gain deeper insight into how prior infections shape the transcriptomic response to MI, we performed single-cell RNA sequencing on infarcted hearts from naïve and MCMV-infected C57BL/6J mice. Five days after MI, the hearts were collected and dissociated into single-cell suspensions for analysis. Both hematopoietic (defined as viable, Calcein^+^ CD45^+^ cells) and non-hematopoietic cells (defined as viable, Calcein^+^ cells CD45^-^ CD31^+^ or CD45^-^ CD31^-^ cells) were sequenced to characterize the gene expression profiles of immune and non-immune cell populations, using a modified protocol from Skelly et al.^28^ (Fig. 4a, S4A). Sample multiplexing using oligo-tagged (hashtag) antibodies was conducted and three independent single-cell sequencing runs were performed. Doublet and low-quality cell exclusion were followed by hashtag demultiplexing and batch correction (Fig. S3). A total of 14,094 leukocytes and 12,844 non-hematopoietic cells were recovered and integrated from the three independent experimental sessions. These included diverse myeloid populations, B cells, and T cells, as well as non-immune cell types such as endothelial cells, fibroblasts, and epithelial cells. The scRNA-seq dataset from Forte et al.^29^ was used as a reference for annotating the different cardiac cellular subsets post MI (Fig 4b, S4B). Our data revealed a higher fraction of B (*Cd79a*) and T (*Cd3e, Trac*) cells in the previously infected mice and a greater proportion of cells composing the epicardium (*Wt1, Dmkn*) in the MCMV+MI group, compared to naive mice (Fig. 4c). As previously shown, these epicardial cells make up the mesothelium covering the heart and are the major source of cardiac fibroblasts during homeostasis^29^. Several cell types, including cardiac macrophages and myofibroblasts, exhibited an overall pro-inflammatory state that was absent in the naive MI group (Fig. 4d, S5). Gene set enrichment analysis (GSEA) identified a significant upregulation of genes associated with TNF signaling, KRAS signaling, allograft rejection, and inflammatory responses in cardiac macrophages and in *Arg1*-expressing monocytes (Fig. S5) of MCMV-exposed mice. Among the upregulated genes were *Il1b*, *Il6*, *Ccl2*, *Cxcl2*, *Cxcr4*, and *Igf1*, reflecting activation of pro-inflammatory and chemotactic pathways as well as signaling processes associated with immune regulation and tissue Myofibroblasts from MCMV+MI mice displayed a similar pattern, with additional responses to interferon-α and -γ, thus indicating involvement in antiviral defense and inflammatory processes.

**Fig. 4.**
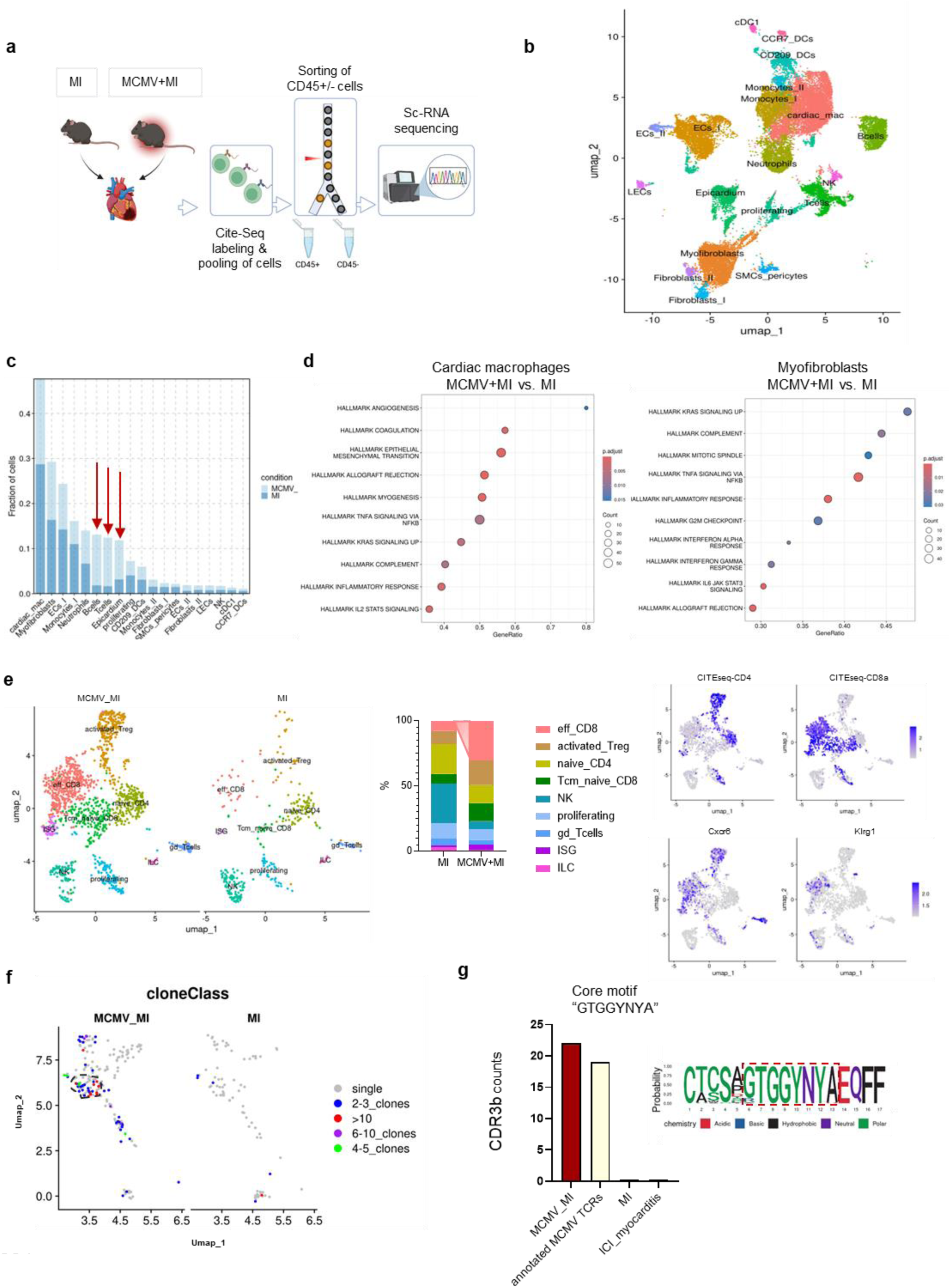
Prior MCMV exposure induces an elevated pro-inflammatory transcriptional profile in the cardiac tissue of infarcted mice. **a.** Experimental outline for the sc-sequencing pipeline. Infarcted hearts from naive (MI) or MCMV-infected (MCMV+MI) mice were collected 5 days after MI and processed to obtain a single-cell suspension. Individual mice were labeled with CITE-seq antibodies and cells were eventually pooled to sort for CD45+ and CD45-CD31^+^ and CD45^-^ CD31^-^ cells, and single-cell sequencing was performed using the 10x Genomics platform. **b.** UMAP plot representation of the sorted CD45+ and CD45-cells (approx.15,000 and 13,000 cells, respectively) from 12-week-old male naive and MCMV-infected mice, 5 days post MI. **c.** Bar graph representation of the cell proportion of MCMV+MI or MI in each annotated cell cluster. **d.** Gene set enrichment analysis of the top upregulated genes from cardiac macrophages and myofibroblasts in MCMV+MI mice compared to naive controls (MI). The gene list was tested against the Hallmark gene sets from the Molecular Signatures Database (MSigDB). Dot size indicates the number of ranked-list genes that overlap with a gene set. Color demotes adjusted p-value from the enrichment test. **e.** UMAP visualization of the subclustered T- and NK-cell subsets for each one of the two conditions, and bar graph quantification of the cell proportions. Feature plots indicate CITE-seq expression for CD4 and CD8, as well as *Cxcr6* and *Klrg1* gene expression. **f.** Feature plots of TCR CDR3 sequences indicating clonal expansion in MCMV+MI or MI conditions overlaid on the subclustered CD4 and CD8^+^ T cells that have a productive TCR sequence. Grey indicates a unique TCR CDR3 sequence (single), blue color indicates a CDR3 sequence overlapping in 2-3 T cells (2-3 clones), green (4-5 clones), violet (6-10 clones), and red (>10 clones) **g.** Barplot of the number of CDR3 sequences that have an overlapping core motif “GTGGYNYA” with annotated MCMV specific TCRs from the McPAS-TCR database^31^. And a sequence logo representing the aligned CDR3β amino acid sequences showing conserved positions and variability within the motif.

T lymphocytes are key players in antiviral immunity and based on our previous data, MCMV-specific T cells can infiltrate and persist in the heart after infection (Fig 1,2), hence we were interested in taking a deeper look into this compartment. We therefore subclustered all T and NK cells, which resulted into a cluster containing activated Tregs (*Icos*, *Ctla4*), effector CD8^+^s (*Cd8a*, *Klrg1*, *Cxcr6*), naive CD4s (*Igfbp4, Ccr7*), T central memory and naive CD8^+^s (*Ly6c2*, *Sidt1*), and natural killer cells (NK, *Ncr1, Klra8, Klrb1c*); a cluster of proliferating cells (*Mki67*) and γδ T cells (*Trdc*, *Trg-C1*); and a cluster with high levels of interferon-stimulated genes (ISG, *Ifit1*, *Isg15*) and group 2 innate lymphoid cells (ILC2, *Gata3*, *Rora*) (Fig. 4e, S4C). We observed an expansion of the eff_ CD8 cluster in the MCMV+MI group and a contraction in the proportion of NK and naive_CD4 cells compared to the naive MI mice. Furthermore, we were able to identify CD4^+^ and CD8^+^ cells by the respective CITE-seq expression and observed an overlapping expression with Cxcr6 and Klrg1 for both cell types. These differences in these specific cell proportions, as well as gene expression, signify both an activated immune system dominated by CD8^+^ T cells and an established protective memory due to viral pre-exposure.

During development, T cells undergo somatic recombination of variable gene segments, resulting in a highly diverse repertoire of receptors that can recognize a vast array of antigens. Each T cell expresses a unique clonotypic T cell receptor (TCR), and this diversity underlies the specificity of T cell antigen recognition. After CMV infection, T cells undergo large clonal expansions during memory inflation, and various specific CMV-related TCRs have previously been annotated. In view of this, we used TCR sequencing paired with single-cell gene expression and identified a subset of T cells that clonally expanded in the infarcted hearts of MCMV-infected mice but not the naive ones (Fig. 4f). To infer the antigen specificity of expanded TCRs, we probed for their overlap with annotated MCMV-responding TCRs from the McPAS-TCR database^30^, a manually curated repository of TCR sequences associated with various pathologies,^30^ including responses to viral infections such as MCMV. Next, we applied the GLIPH2 algorithm^31^, which models TCR antigen specificity based on sequence similarity of the complementarity-determining region 3 (CDR3). GLIPH2 is critical for determining antigen specificity because it identifies conserved core motifs of short amino acid sequences within the CDR3 region of the TCR that directly contacts the antigenic peptide presented by the MHC molecule. To control for random motif enrichment, the experimental TCR dataset is compared against a reference repertoire of 78,116 non-disease-associated mouse TCRs^31,32^. Using this approach, we described a core CDR3 motif, “GTGGYNYA”, that exclusively overlapped between the cardiac-expanded TCRs from MCMV infected mice and the MCMV-specific TCRs annotated in previous studies^30^. This core motif was not found in CDR3 sequences from non-infected MI operated mice, nor in TCRs related to cardiac antigens^33^ (Fig. 4g, S5D). Overall, these observations confirm the expansion of MCMV-specific T cells in myocardium poised with a tissue-resident phenotype.

### Presence of CMV-responsive T cells in the cardiac tissue of MI patients links worse healing after MI with CMV IgG titers

In clinical cohorts, CMV has increasingly been linked to adverse cardiovascular outcomes, with studies showing an association between CMV seropositivity and a higher prevalence of coronary heart disease^9^ as well as age-related pathologies including cardiovascular aging^18^. Our findings from mouse models so far indicate that CMV infection prior to a cardiac ischemic event resulted in the accumulation of CMV-reactive T cells with a cardiac-resident phenotype, an altered inflammatory landscape during the post-MI response, and impaired cardiac healing. To interrogate a possible correlation between CMV infection and post-MI outcomes in patients, we employed a retrospective analysis from the Etiology, Titre-Course, and Survival (ETiCS) Study^34^. Previously, patients from the ETiCS cohort were stratified as either “good healers” based on improved left ventricular ejection fraction (ΔLVEF) > 13% at the 12-month follow-up (FUP) time-point, compared to index hospitalization, or “poor healers” if their ΔLVEF < 13% (Fig. 5a) ^35^. The ΔLVEF > 13% cutoff was determined, by Bulluck et al. as the minimal effect size required for LVEF improvement following an acute STEMI event.^36^ While good and poor healers had no observable difference in CMV seropositivity, poor healers exhibited higher CMV IgG titers compared to good healers (Fig. 5b). We next made use of publicly available TCR sequencing analyses of blood samples from good and poor healers^35^ and computed the overlaps between highly expanded TCRs in good or poor healing patients and human CMV-responding TCRs annotated in the McPAS-TCR database. By computing the Jaccard similarity score between CMV-expanded TCRs and those expanded in good or poor healers, which is an index reflecting the proportion of shared TCR sequences relative to the total number of unique sequences, we found that CMV-expanded TCRs exhibited more similarity with the poor healer group (Fig. 5c). In addition to CMV, we also curated EBV-annotated TCRs from the McPAS-TCR database. We detected EBV-responding TCRs in the donor blood samples, a result we expected given the high prevalence of EBV in the general population^37^. However, in contrast to CMV, no preferential expansion of EBV-annotated TCRs was observed in the poor healer group (Fig. S6). Next, we performed TCR sequencing on cardiac tissue obtained from autopsies of deceased MI patients and biobanked at the Medical University of Graz (Fig. 5d). CMV-annotated TCRs overlapped with expanded TCRs from cardiac samples, a result that indicates CMV-reactive T cells are present in the myocardium. Interestingly, a higher clonal overlap of CMV TCRs was observed between cardiac tissue and blood samples from poor healers compared to good healers (Fig. 5e). Additionally, TCRs reactive to defined CMV epitopes exhibited a distinct overlapping pattern in TCRs from poor healers and those from cardiac tissue. TCRs reactive to the CMV epitopes pp50, IE-1, and UL29/28 showed clonal overlap among the TCR repertoire of cardiac samples and blood samples from the poor healers group, but not the good healers group (Fig. 5f). By similarly analyzing TCRs reactive to defined EBV epitopes, we noted a distinct pattern of epitope-specific TCRs in the blood samples between the good and poor healer groups. However, cardiac tissue samples had less expansion of EBV epitope-specific TCRs, and there was minimal overlap between EBV-specific TCRs in cardiac tissue and those found in the blood of poor healers (Fig. S6). These findings establish a clear association between the presence of CMV-reactive T cells in the heart and outcomes after MI, a connection corroborated by our results from experimental mouse models. Our findings thus emphasize the pivotal role of the immune response in influencing post-myocardial infarction healing outcomes in CMV-positive patients, underscoring significant clinical implications for patient stratification and the development of innovative immune-targeted therapies.

**Fig. 5.**
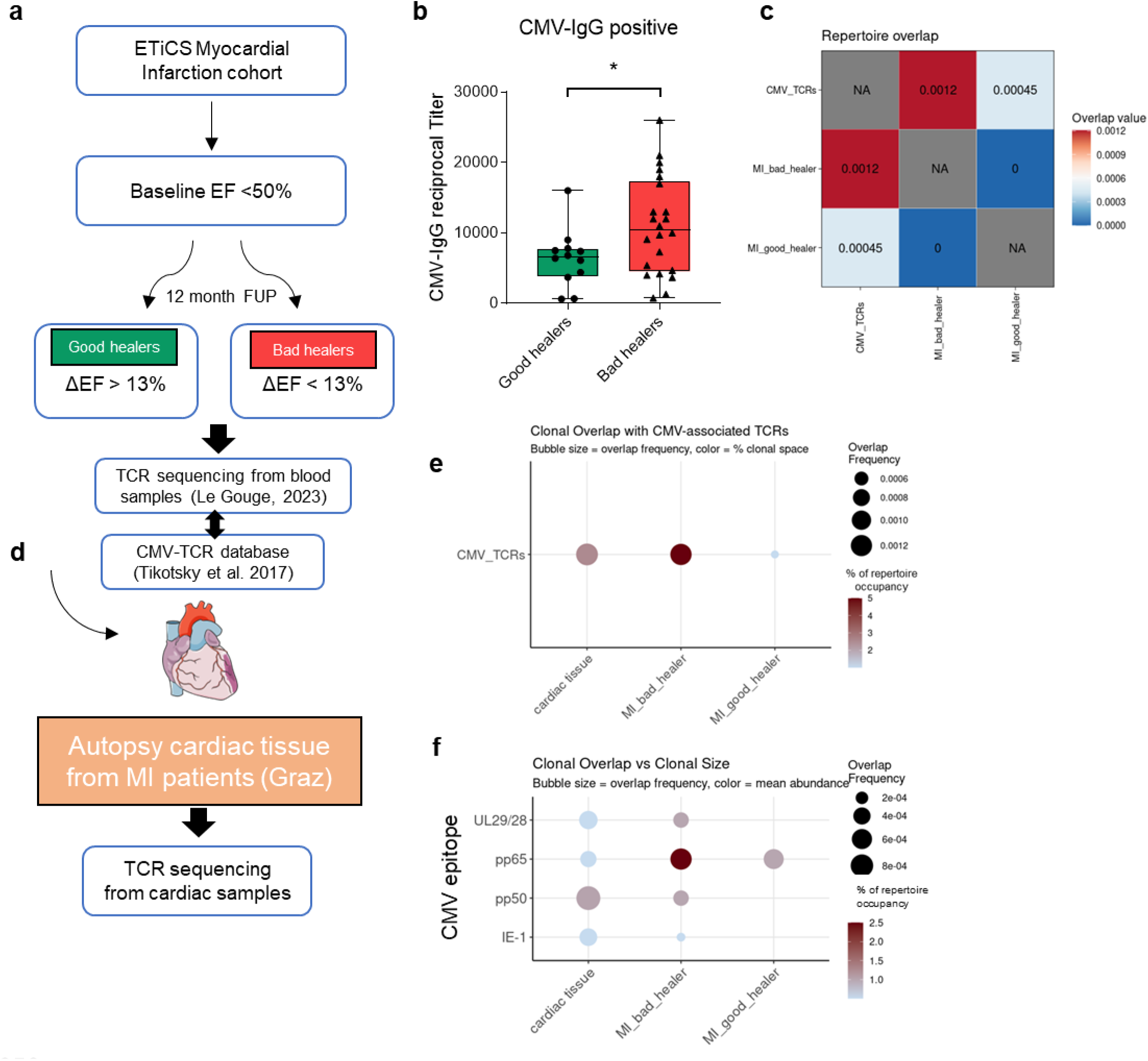
Presence of CMV-responsive T cells in the cardiac tissue of MI patients links worse healing after MI with CMV IgG titers. **a.** Study design and ΔLVEF-based stratification of MI patients from the ETiCS cohort into good or poor healers. **b.** Frequency of MI patients who are CMV-IgG seropositive or seronegative (left), and CMV-IgG reciprocal titers from the seropositive MI patients categorized as good or poor healers. Statistics were calculated using unpaired Student t-test; p value is numerically indicated. **c.** Heatmap of Jaccard similarity index based on exact CDR3b sequence matches between CMV-annotated TCRs^30^and TCR repertoires from blood samples of poor or good post-MI healing groups. Color intensity reflects the degree of clonal similarity, with red indicating higher overlap. **d.** TCR sequencing from cardiac autopsies of acute MI samples and integration with TCRs from blood samples of the ETiCs cohort. **e.** Dotplots of Jaccard similarity index between CMV-annotated TCRs, TCR repertoires from blood samples of poor or good healers, and cardiac autopsies. **f.** Dotplot of Jaccard similarity index between the same groups comparing their specificity to defined CMV epitopes. Dot size reflects the degree of clonal similarity, and color intensity indicates the degree of clonal expansion of CMV TCRs within each analyzed group.

## Discussion

In this study, we explored how infection with CMV, a widespread herpesvirus, affects cardiac homeostasis both in steady-state and following acute MI. In latently infected mice we observed a notable enrichment of cardiac leukocytes, particularly tissue-resident T cells and monocyte-derived macrophages, accompanied by a distinct proinflammatory transcriptional profile. After MI, these differences became even more pronounced. Multiple cardiac cell populations, including fibroblasts and macrophages, exhibited an amplified inflammatory state, and T cells with a tissue-infiltrating effector phenotype were markedly more abundant in latently infected mice compared to naive controls. These immunological changes were associated with signs of adverse cardiac remodeling during both the acute and chronic phases of MI, as evidenced by increased diastolic volumes in latently infected mice. Finally, we identified a correlation between the presence of CMV-responsive T cells in human cardiac tissue and poorer clinical outcomes after MI. While the link between CMV seropositivity and cardiovascular risk has been recognized for nearly four decades, as far as we know, this is the first experimental study to mechanistically address the interplay among CMV infection, myocardial immune cells, and post-MI repair.

Cardiovascular diseases have been extensively studied in murine models. Although laboratory mice are considered to closely mimic human cardiac function and tissue composition, their immune systems are not shaped by natural pathogen exposure, which can have a major influence on tissue homeostasis^38^. Latent CMV has been shown to change the leukocyte composition of the lungs, making them more resistant to bacterial^39^ and viral^40^ challenges. Furthermore, transcriptomic analyses of liver, spleen, and brain tissue of mice latently infected with a herpesvirus closely related to CMV showed a clear proinflammatory change in gene expression patterns^41,42^. However, until now, a comprehensive analysis of the leukocyte compartment within cardiac tissue following CMV exposure has not been conducted. Latent MCMV infection produced distinct transcriptional and metabolic profiles in the heart, compared to non-infected animals. More specifically, during viral latency, we observed a downregulation of oxidative phosphorylation and respiratory pathways along with upregulated interferon responses. This inverse correlation has been previously reported in various contexts and could indicate a role of interferon in reshaping the cardiac metabolic homeostasis during latent infections^14^. Our data are consistent with the idea that long-term persisting resident T cells could maintain this altered state in the myocardium during latency.

Our findings demonstrate pronounced leukocyte infiltration that began during the acute phase of infection and persisted above baseline levels even during MCMV latency. Among these infiltrating leukocytes, we particularly noted a population of CD8^+^ T cells that recognize an epitope derived from m139 MCMV protein and had all the phenotypical features of tissue-resident memory T cells (negative for intravascular CD45 staining and positive for CD69, CXCR6, CD38). Tissue-resident T cells have been extensively characterized in various organs but seldomly identified in cardiac tissue. A recent study by Kalinoski et al. noted a population of cardiac myosin-specific tissue resident memory (Trm) cells which were associated with an increased susceptibility of mice to develop immune checkpoint-induced myocarditis^43^. Trm T cells were also found in the epicardial tissue of patients with atrial fibrillation^44^. Nevertheless, to the best of our knowledge, our study is the first to define a population of virus-specific cardiac T cells with a Trm phenotype.

CMV has been identified in cardiac tissue through both post-mortem human studies^22^ and experimental mouse models^24^. Consistent with these observations, we noted active viral replication within cardiac tissue for up to nine days following infection (Fig. 1c, d). Several epidemiological and other studies have linked latent CMV infection to poorer outcomes after cardiovascular events^18,21^. Supporting this association, our analysis of a cohort of patients with MI revealed that elevated anti-CMV antibody titers correlated with diminished cardiac function 12 months after the initial event. Moreover, TCR sequencing uncovered a notable overlap between CMV-annotated TCRs and those expanded in patients who showed inadequate improvement in left ventricular ejection fraction post-MI, an overlap not found with EBV-annotated TCRs. These dynamics further support the connection between CMV infection and adverse cardiac outcomes. Our animal studies reinforced these clinical observations as echocardiography and cMRI assessments, performed 5 and 56 days after MI respectively, determined that MCMV^+^ mice had worse adverse cardiac remodeling compared to infarcted controls maintained under specific pathogen-free conditions.

Perturbing a steady-state condition after experimental MI exacerbated the changes in the cardiac immune cell composition of MCMV^+^ versus MCMV^-^ mice. This shift was primarily driven by elevated CD8^+^ T cell and monocyte/macrophage influx. Furthermore, our data revealed a pro-inflammatory gene expression signature broadly distributed across various cell types, ranging from immune cells like macrophages to non-immune cells like myofibroblasts, in the infected and infarcted group. The latter are known to peak between day 3 and day 7 post MI^29^; thus, heightened inflammatory responses by such an abundant cell population during this time period are likely to significantly influence scar formation and tissue remodeling.

The profoundly varied inflammatory milieu in the hearts of MCMV^+^ hosts compared to MCMV^-^ controls could explain the differences observed in clinical outcomes and cardiac remodeling after MI. T cells, especially those harboring the Trm phenotype, were considerably enriched in CMV^+^ mice. In line with our FACS and RNA-sequencing findings, our TCR-sequencing data revealed a clonal expansion of MCMV-specific CD8^+^ T cells that reside in the myocardium, a shift that suggests the heightened inflammatory state might result from these virus-specific cells first infiltrating and then locally replicating in cardiac tissue. Several studies have shown that persistent T cell stimulation in cardiac tissue can lead to adverse effects after MI. Indeed, activated T cells secreting IFN-γ, IL-17, and TNF have been found to promote fibrosis development and adverse cardiac remodeling during the chronic phase of MI^45^. Furthermore, Ilatovskaya et. al reported that CD8^+^ T cell-deficient mice have improved survival after MI despite having substantially elevated risk for cardiac rupture during its early stages^46^. This increased cardiac rupture was associated with inadequate cross-linking of collagen fibers, a phenomenon which highlights the effect that T cells could have on cardiac fibroblasts. Our single-cell RNA sequencing data uncovered a distinct inflammatory phenotype, characterized by interferon and TNF gene signatures, among cardiac myofibroblasts in MCMV^+^ mice. Cardiac fibroblasts are extensively regulated by inflammatory cues received from their environment^47^ and an inflammatory milieu generated by latent CMV infection could lead to pathological collagen deposition, ultimately resulting in adverse remodeling.

Our flow cytometry and single-cell RNA sequencing analyses also showed pronounced alterations in macrophage composition and phenotype within the hearts of MCMV^+^ mice. Notably, viral infection was associated with a marked shift toward a CCR2^+^ macrophage population, and transcriptomic data showed an inflammatory signature among cardiac macrophages. CCR2^+^ macrophages are derived from hematopoietic progenitors and are replenished by circulating monocytes. Upon myocardial damage, these macrophages produce high levels of inflammatory cytokines and recruit neutrophiles and monocytes, hereby inducing generation of oxidative products in cardiac tissue^48^. This macrophage population is associated with excessive inflammation and adverse cardiac remodeling, as blocking CCR2 or selectively depleting CCR2^+^ macrophages resulted in improved cardiac function and smaller LV sizes after experimental MI^49^. It is therefore plausible that MCMV infection primes the cardiac macrophage population toward a heightened proinflammatory state, which, following MI, may exacerbate inflammation and ultimately contribute to adverse cardiac remodeling.

To conclude, here we present an in-depth molecular and cellular analysis of changes induced in cardiac tissue by a common herpesvirus infection. We experimentally explored and validated long-standing epidemiological data linking CMV infection with worse cardiac outcomes after cardiovascular events. These observations should encourage further investigations into cardiac immune responses following cardiotropic infections and highlight pre-exposure to CMV as a predictive factor for tissue remodeling and healing outcomes post MI.

## Funding

GCR is supported by the German Research Foundation (DFG grant number 517001338), and lead projects integrated in the Collaborative Research Centre 1525 ‘Cardio-Immune Interfaces’ (funded by the German Research foundation grant number 453989101). SJ, IB and MS are supported by the grant “Strengthening the capacity of CerVirVac for research in virus immunology and vaccinology” (KK.01.1.1.01.0006) granted to the Scientific Centre of Excellence for Virus Immunology and Vaccines and co-financed by the European Regional Development Fund. MS is supported by grants “uniri-mladi-biomed-22-44”, “uniri-mladi-biomed-23-53” and “uniri-mz-25-63” financed by University of Rijeka. This work was supported by the European Research Area Network—Cardiovascular Diseases [ERANET-CVD JCT2018, AIR-MI Consortium to GCR (01KL1902), PPR (4168-B) and EMF (ANR-18-ECVD-0001)].

## Data Availability

The detailed methods and additional figures are available in the Supplemental Material. Transcriptomic datasets from this study have been archived in NCBI's Gene Expression Omnibus (GEO) database (GSE309570 and GSE309706).

## Acknowledgements

We thank Lisa Popiolkowski, Dijana Rumora, Ante Miše, Mihaela Gašparević, Cristina Paulović, Antonija Šarlija, Andrea Leupold, and Elena Vogel for their skillful technical assistance. We also thank the Core Unit SysMed at the University of Würzburg for excellent technical support, RNA-seq data generation, and analysis. We also thank the NIH Tetramer Core Facility for providing m139, p79 and NP tetramers. The figures were prepared with the help of SMART Servier Medical Art and Biorender.

## Author contributions

Conceptualization: IB, GG, GCR; Methodology: ED, MS, DA, MCB, JM, BL, KK, MT, KL, EMF, PRR, TK, AES, SJ, UH, SF, IB, GG, GCR; Investigation: ED, MS, DA, MCB, JM, BL, VS, LR, MT, TK, KL; Funding acquisition: PPR, IB, GG, GCR; Project administration: IB, GG, GCR; Supervision: IB, GG, GCR; Writing – original draft: ED, MS, DA, GCR; Writing – review & editing: all.

## Conflict of Interests

None.

## Data and materials availability

The murine bulk RNA sequencing data and single cell RNA sequencing data from this study are deposited in NCBI’s Gene Expression Omnibus ^42^ database (GSE309570 and GSE309706). Anonymized patient sequencing data will be made available upon reasonable request.

## List of nonstandard abbreviations

CDR3: Complementarity-determining region 3
cMRI: Cardiac magnetic resonance imaging
CMV: Cytomegalovirus
EBV: Epstein–Barr virus
GLIPH: Grouping of lymphocyte interactions by paratope hotspots
IFN-γ: Interferon Gamma
IL-6: Interleukin 6
LVEF: Left ventricular ejection fraction
MI: Myocardial infarction
MHC: Major histocompatibility complex class
RTM: Resident tissue macrophage
STEMI: ST-elevation myocardial infarction
TCR: T cell receptor
TCR-M: Myosin-specific T cell
TNF: Tumor necrosis factor

## Notes

### Competing Interest Statement

The authors have declared no competing interest.

### Author Declarations

Animal procedures were approved by the local authorities (Regierung von Unterfranken and Animal Welfare Committee at the University of Rijeka) and conformed to the guidelines from Directive 2010/63/EU of the European Parliament on the protection of animals used for scientific purposes. The use of human tissue samples from Graz is conformed with legal and institutional requirements and was approved by the ethics committee of the Medical University of Graz (31-288 ex 18/19). The ETiCS study from Würzburg was approved by the ethics commission of the University of Würzburg (186/09) and met the criteria of the Declaration of Helsinki. All patients provided written informed consent.

